# The SARS-CoV-2 T-cell immunity is directed against the spike, membrane, and nucleocapsid protein and associated with COVID 19 severity

**DOI:** 10.1101/2020.05.13.20100636

**Authors:** Constantin J. Thieme, Moritz Anft, Krystallenia Paniskaki, Arturo Blazquez-Navarro, Adrian Doevelaar, Felix S. Seibert, Bodo Hoelzer, Margarethe Justine Konik, Thorsten Brenner, Clemens Tempfer, Carsten Watzl, Sebastian Dolff, Ulf Dittmer, Timm H. Westhoff, Oliver Witzke, Ulrik Stervbo, Toralf Roch, Nina Babel

## Abstract

Identification of immunogenic targets of SARS-CoV-2 is crucial for monitoring of antiviral immunity and vaccine design. Currently, mainly anti-spike (S)-protein adaptive immunity is investigated. However, also the nucleocapsid (N)- and membrane (M)-proteins should be considered as diagnostic and prophylactic targets.

The aim of our study was to explore and compare the immunogenicity of SARS-CoV-2 S-, M- and N-proteins in context of different COVID-19 manifestations. Analyzing a cohort of COVID-19 patients with moderate, severe, and critical disease severity, we show that overlapping peptide pools (OPP) of all three proteins can activate SARS-CoV-2-reactive T-cells with a stronger response of CD4^+^ compared to CD8^+^ T-cells. Although interindividual variations for the three proteins were observed, M-protein induced the highest frequencies of CD4^+^ T-cells, suggesting its relevance as diagnostic and vaccination target. Importantly, patients with critical COVID-19 demonstrated the strongest T-cell response, including the highest frequencies of cytokine-producing bi- and trifunctional T-cells, for all three proteins. Although the higher magnitude and superior functionality of SARS-CoV-2-reactive T-cells in critical patients can also be a result of a stronger immunogenicity provided by severe infection, it disproves the hypothesis of insufficient SARS-CoV-2-reactive immunity in critical COVID-19. To this end, activation of effector T-cells with differentiated memory phenotype found in our study could cause hyper-reactive response in critical cases leading to immunopathogenesis. Conclusively, since the S-, M-, and N-proteins induce T-cell responses with individual differences, all three proteins should be evaluated for diagnostics and therapeutic strategies to avoid underestimation of cellular immunity and to deepen our understanding of COVID-19 immunity.

## Main text

Clearance of viral pathogens requires an effective T cell response directed against protein antigens expressed by the virus^1^. The T cell response against the severe acute respiratory syndrome-related coronavirus (SARS-CoV)-2 virus, which causes the ongoing pandemic, is presumably initiated by respiratory professional antigen presenting cells (APC) that can engulf viral antigens as shown for the 2002/03 SARS-CoV^2^. The activated T cells can migrate to the site of infection, where they facilitate viral clearance, but can also contribute to immune pathogenesis. There is mounting evidence that the latter is the major reason for critical COVID-19 disease manifestations^3,4^.

The SARS-CoV-2 contains four structural proteins: the spike glycoprotein (S), the envelope (E) protein, the membrane (M) protein and the nucleocapsid (N) protein^5^. The S-protein mediates host cell entry by binding to the angiotensin-converting enzyme 2 (ACE2)^6^. Due to its surface-exposure and crucial role for infecting the host cell, the S-protein is an attractive therapeutic target, for instance for antibodies that block the S/ACE2 interaction. In fact, it was shown that patients recovered from COVID-19 developed virus neutralizing anti-S immunoglobulin (Ig) titers^7^. Given the requirement for T cell help in generation of high affinity IgG antibodies, this finding indicates that S-protein reactive T cell immunity was formed in those patients^8^,^9^. Accordingly, very recent studies identified SARS-CoV-2 S-protein reactive T cell responses in patients suffering from moderate, severe, and critical COVID-19^4^,^10^. Furthermore, it was shown that the amount of SARS-Co-V2 reactive T cells increased with disease progression^11^. Besides the S-protein, also the N- and M-proteins were suggested as potential targets for diagnostic and prophylactic approaches^3,5^. In fact, B cell responses against the N-protein seemed to be the first to arise 4–8 days after symptom onset for the 2002/03 SARS-CoV infection^12,13^, which indicates that also N-reactive T cell response are prevalent during this timeframe. However, data on T cell responses towards the N- and M-proteins for the pandemic SARS-CoV-2 infection are currently not available. For this reason, we sought to identify, characterize, and compare S-, M-, and N-reactive T cell responses in COVID-19 patients with different clinical manifestation.

We analyzed 57 blood samples drawn at different time points after hospital admission of a cohort of 28 COVID-19 patients with moderate (28 samples), severe (16 samples), and critical (13 samples) disease manifestation (Table S1). The samples were grouped according to the COVID-19 severity at the sampling time into moderate, severe, critical manifestation using the German Robert-Koch-Institute symptom classification as previously described^4^. In agreement with other studies^14^, we observed significantly more males within critical COVID-19 patients compared to the moderate and severe cases. However, bivariate regression analysis revealed no significant influence of gender on the main findings when comparing different COVID-19 severity (Table S2). There were no significant differences in sampling time with respect to the PCR-testing or hospitalization between the groups (Table S3).

By stimulation with S-, M-, or N- overlapping peptide pools (OPP) (Fig. S1), we could show that all three proteins have the capacity to induce SARS-CoV-2-reactive CD4^+^ and CD8^+^ T cells (Fig. 1a). Overall, we detected a CD4 +T cell response in 54 out of 57 and a CD8+ T cell response in 44 out of 57 of the patient samples against at least one of the SARS-CoV-2 proteins. However, none of the proteins was able to induce CD4^+^ or CD8^+^ T-cell response in all 54 and 44 positive samples, respectively. Within the 54 responding samples, M-protein OPP induced a detectable CD4^+^ T cell response in the highest number of samples (M=45, N=39, S=44), whereas for the 44 responding samples within CD8+ subsets, the S protein OPP was dominant (M=30, S=36, N=30) (Fig. 1b-c). A similar observation was made for the middle eastern respiratory syndrome corona virus (MERS)-virus, where a higher S-protein induced CD8^+^ T cell and higher M-protein induced CD4^+^ T cells response were described^15^. Although the immunogenicity of the three proteins and its role for the clinical course require follow up investigations, our data advocate that measuring T cell responses induced by all three proteins is of high relevance when studying SARS-CoV-2-reactive T cells.

**Fig 1:**
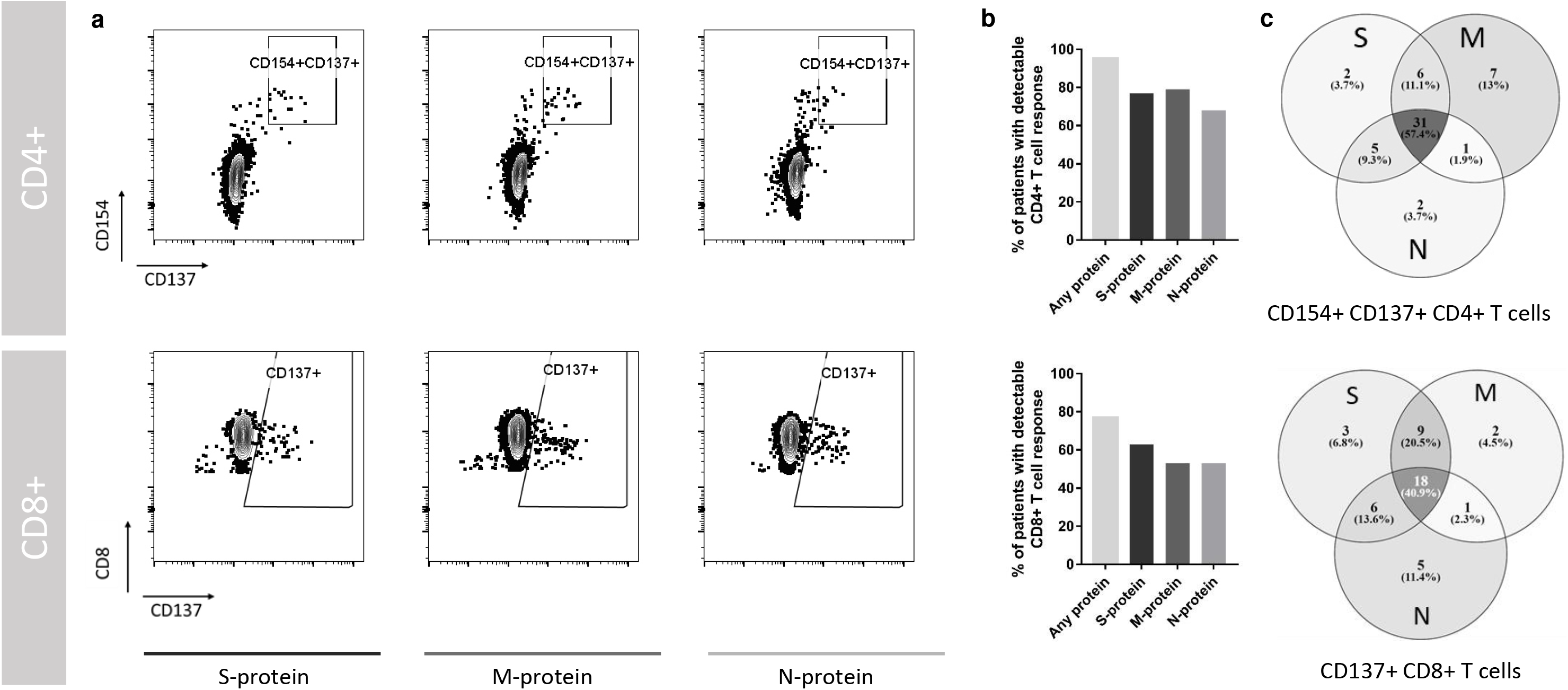
SARS-CoV-2–reactive T cells are induced by the S-, M- and N- protein with interindividual pattern. Peripheral blood mononuclear cells isolated from 57 blood samples of a total of 28 COVID-19 patients with moderate, severe or critical disease were stimulated for 16 hours with S-, M-, or N-protein OPP. Antigen-reactive T cells were determined by flow cytometry and identified according to the gating strategy presented in supplementary figure S2. **a)** Representative plots of CD4^+^ T cells and CD8^+^ T cells after stimulation with S-, M-, and N-protein overlapping peptide pools for 16h. Antigen-reactive CD4^+^ T cells were identified by CD154 and CD137 expression and antigen-reactive CD8^+^ T cells by CD137 expression. **b)** Frequency of patient samples with detectable CD4^+^ (top) and CD8^+^ (bottom) T cell responses after stimulation with S-, M-, or N-protein (total of 57 samples of 28 patients). **c)** Venn diagrams of 57 patient samples with detectable SARS–Cov–2-reactive CD4^+^ or CD8^+^ T cells after stimulation with S-, M- or N-protein. Antigen-specific responses above 0.001% of CD4^+^ or CD8^+^ T cells were considered as response. 54 samples within CD4^+^ T-cells and 44 samples within CD8^+^ T-cells showed T cell reactivity towards at least one of the tested SARS– CoV–2S, M, and N-proteins

Comparing the magnitude of response against the three proteins, we also found that the M-protein OPP induced the highest frequencies of reactive CD4^+^ T cells (Fig. 2a). Compared to the S- and N-reactive CD4^+^ T cells we found a consistent trend of higher frequencies of M-reactive CD4^+^ T cells expressing cytokines and effector molecules such as interleukin (IL)2, interferon γ (IFNγ), tumor necrosis factor α (TNFα), and granzyme B (GrzB). N-protein induced the lowest responses in comparison to the other proteins (Fig. 2b-f, Fig. S2). Despite the differences in magnitude, there was a high correlation between the frequencies of S-, N-, and M-reactive CD4^+^ T cells (Fig. 2g-i).

**Fig 2:**
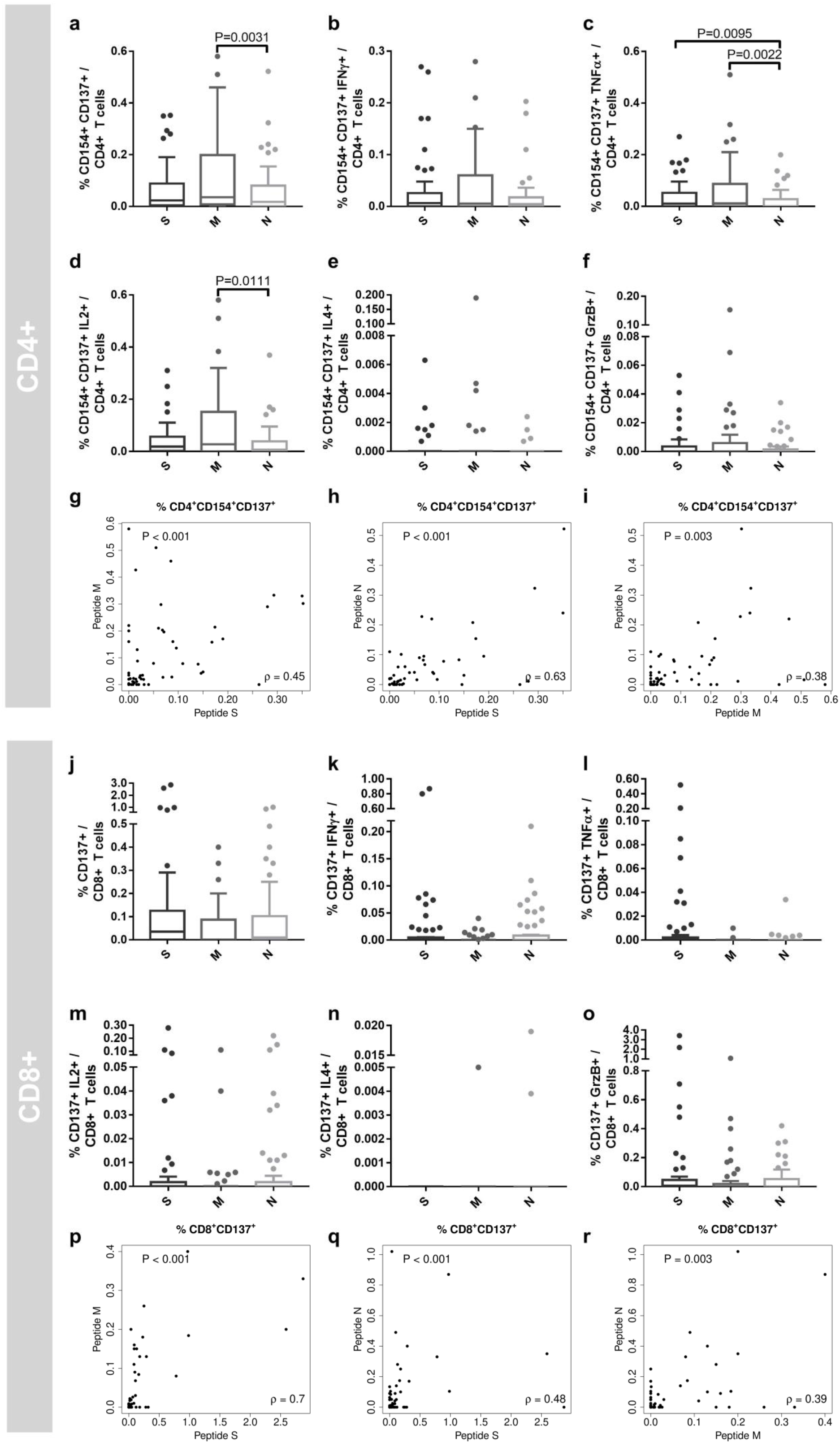
SARS-CoV-2 reactive T cells show a higher M-protein reactivity in CD4^+^ T cells and S- and N-protein reactivity in CD8^+^ T cells. 57 blood samples of a total of 28 COVID-19 patients were drawn at one or at multiple time points within one week after diagnosis. Peripheral blood mononuclear cells were stimulated for 16h with S-, M-, or N-protein overlapping peptide pools. The gating strategy is presented in Fig.S2. **a-f**) Frequencies of (a) CD154^+^ CD137^+^ CD4^+^ T cells (antigen-specific CD4^+^ T cells), (b) interferon γ (IFNγ)–, (c) tumor necrosis factor α (TNFα)–, (d) interleukin (IL) 2–, (e) IL4–, and (f) granzyme B (GrzB)-producing antigen-specific CD4^+^ T cells. Statistical analysis was performed with Friedman test for non-parametric data and Dunn’s multiple comparison test. Whiskers were calculated with the Tukey method. **g-i**) Correlation of M-, N- and S-protein OPP reactive (CD154+ CD137+) CD4+ T cells. Calculation was performed with Spearman’s rank correlation coefficient. **j-l)** Frequencies of (j) CD137^+^ CD8^+^ T cells (antigen-specific CD8^+^ T cells), (k) IFNγ, (l) TNFα-, (m) IL2, (n) IL4, and (o) GrzB-producing antigen-specific CD8^+^ T cells. Statistical analysis was performed with Friedman test for non-parametric data and Dunn’s multiple comparison test. Whiskers were calculated with the Tukey method. **p-r**) Correlation of M-, N- and S-protein reactive (CD137+) CD8+ T cells. Calculation was performed with Spearman’s rank correlation coefficient.

Interestingly, the pattern observed for S-, N-, and M-protein reactivity of CD4^+^ T cells were not found in CD8^+^ T cells (Fig. 2j-o). In fact, a tendency of higher frequencies of S- or N-protein reactive CD8^+^ T cells compared to M-protein was observed, but without reaching statistical significance after correction for multiple testing. Similar to CD4^+^ restricted immunity, we observed a high correlation in frequencies of CD8^+^ T cells reactive to S-, N-, and M-proteins (Fig 2p-r).

The exact role of SARS-CoV-2-reactive T-cell immunity for COVID-19 progression is currently unknown. We therefore investigated differences in the T cell immunity between moderate, severe and critical COVID-19 patients. A defective switch between innate and adaptive immunity has been described to differentiate patients with favorable and unfavorable outcome after SARS-CoV infection in previous studies^16^. Surprisingly, and in contrast to the endemic SARS-CoV infection, we detected the highest magnitude of CD4^+^ and CD8^+^ T cells reactive to S-, M-, and N-proteins in critical COVID-19 (Fig. 3).

**Fig 3:**
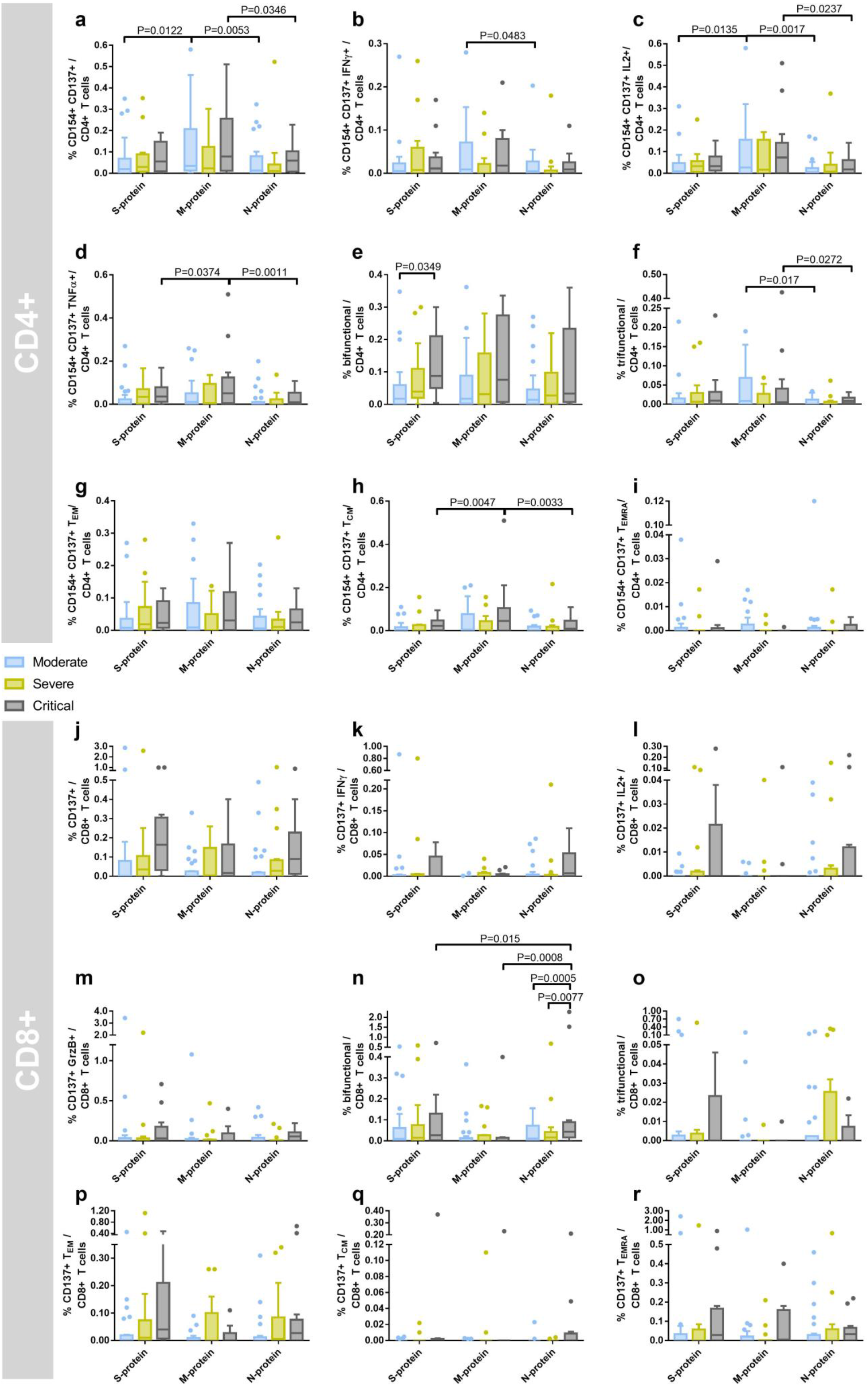
Critical COVID-19 patients mount a strong SARS-CoV-2 S-, M-, and N-protein-reactive CD4^+^ and CD8^+^ T cell response. 57 blood samples of a total of 28 COVID-19 patients were drawn at one or at multiple time points within one week after diagnosis. COVID-19 severity was assessed at the time of sampling as per the guidelines of the German Robert-Koch-Institute and samples grouped accordingly (n=28 moderate, n=16 severe and n=13 critical samples). Peripheral blood mononuclear cells were stimulated for 16h with S-, M-, or N-protein overlapping peptide pools and analyzed by flow cytometry. The gating strategy to identify SARS-CoV-2 S-, M-, or N-protein reactive T cells is presented in Fig. S2 **a-d**) Frequencies of (a) CD154^+^ CD137^+^ CD4^+^ T cells (antigen-specific CD4^+^ T cells), (b) interferon γ (IFNγ), (c) interleukin (IL) 2-, and (d) tumor necrosis factor α (TNFα)producing antigen-specific CD4^+^ T cells. **e-f**) Frequencies of polyfunctional CD4^+^ T cells. (e) Bifunctional and (f) trifunctional CD4^+^ T cells were analyzed by Boolean gating of IL2-, IFNγ, TNFα IL4-, and GrzB-production. A detailed composition of bi- and trifunctional cells is presented in Fig. S3. **g-i)** Frequencies of antigen-specific CD4^+^ (g) T_EM_, (h) T_CM_, and (i) T_EMRA_ cells. **j-m**) Frequencies of (j) CD137^+^ CD8^+^ T cells (antigen-specific CD4^+^ T cells), (k) _IFNγ_, (l) IL2-, (m) GrzB-producing antigen-specific CD8^+^ T cells. **n-o**) Frequencies of polyfunctional CD8^+^ T cells. (n) Bifunctional and (o) trifunctional CD4^+^ T cells were analyzed by Boolean gating of IL2-, IFN_γ_, TNFα IL4-, and GrzB-production. Composition of bi- and trifunctional cells is presented in S3. **p-r**) Frequencies of antigen-specific CD8^+^ (g) T_EM_, (h) T_CM_, and (i) T_EMRA_ cells. Statistical comparisons were done with two-way repeated measurement ANOVA and Tukey’s multiple comparison test. Whiskers were calculated with the Tukey method.

Examining a limited number of subjectively selected functions of virus-reactive T cells may generate distorted and incomplete interpretation of the function and phenotype of these cells^17^. Polyfunctional T cells, that express more than one cytokine or effector molecule, have been described as a hallmark of protective immunity in viral infections^17–19^. Addressing this point, we analyzed the IFNγ, TNFα, IL-2, and IL-4 cytokines as well as the effector molecule GrzB expression in parallel to differentiation stage phenotyping. Of interest, not only the quantity but also the functionality of T-cell immunity was superior in patients with critical COVID-19 severity. Polyfunctional T cells showed higher frequencies in critical COVID-19 patients compared to moderate and severe cases (Fig. 3e,f,n,o). Antigen-reactive IFNγ, IL2, and TNFα-producing CD4^+^ T cells constituted over 50%of trifunctional CD4^+^ T cells, while cytotoxic GrzB-producing IFNγ-and TNFα-expressing CD4^+^ T cells constituted 25% of trifunctional CD4^+^ T cells (Fig. S3b). The cytokine and effector molecule expression of bifunctional CD4^+^ T cells was also dominated by IFNγ, TNFα, IL2, and to a lesser extend GrzB (Fig. S3c). As expected, the majority of polyfunctional CD8^+^ T cells produced the cytotoxic effector molecule GrzB, most commonly in combination with IFNγ and TNFα (Fig. S3b-c). Despite the higher frequencies of single-, bi-, and trifunctional CD4^+^ and CD8^+^ T cells found in most T-cell subsets in critical cases, statistical significant differences were observed only for few subsets. With respect to the relatively low sample numbers and multiple correction testing, the lack of statistical significant differences is not surprising. As described above, SARS-CoV-2-reactive CD4^+^ T cells in our study were predominantly of TH1 phenotype. A T_H_2 dominated response was associated with immunopathology and – eosinophil infiltration after vaccination with N-protein expressing vaccinia virus in a mouse SARS-CoV model^20^,^21^. However, we have no indication of ongoing T_H_2 response in critical patients since only very few IL4-producing T cells were observed in all samples (Fig. 2e-n).

In line with data showing an association between polyfunctionality and the stage of phenotypic differentiation17, we observed higher frequencies of CD8^+^ T cells with effector memory (T_EM_)/T_EMRA_ phenotype in critical COVID-19 patients compared to moderate and severe cases (Fig S4). Importantly, the presence of S-, N-, and M-reactive T-cells with an advanced differentiation phenotype early after diagnosis in our study indicates pre-existing cellular immunity as demonstrated in a recent study^10^.

Taken together, our data provide a comprehensive characterization of the T cell response against S-, M- and N- SARS-CoV-2 proteins. To our best knowledge, this is the first study demonstrating immunogenicity of M- and N-proteins. The reactivity demonstrates individual pattern, indicating that all three proteins should be considered in cellular monitoring to avoid underestimation. Moreover, our findings suggest new potential targets of humoral immunity. Considering the role of CD4^+^ T helper cells for antibody generation and the higher CD4^+^ T-cell response against M-protein, our results highlight the M-protein as an additional target for antibody monitoring and vaccine development.

Importantly, patients with critical COVID-19 demonstrated higher frequencies of CD4^+^ and CD8^+^ T-cells reactive to S-, M, and N-OPP. Although the direct antiviral capacity has to be proven in future studies, the polyfunctionality of the cells might confirm their antiviral potential. To this end, the presence of IFNγ and TNFα coproducing CD4^+^ and CD8^+^ T cells indicating an effector/memory phenotype and long-term protection was also shown for the 2002/03 SARS-CoV^22^,^23^. Considering possible bias in our findings, time point of sample collection and multiple samples per patients might influence our results. However, analyzing only one sample per patients using the last visit sample for an optimized timing match, we did not detect relevant differences to the analysis of all available samples (Fig. S4). We also did not find any significant differences in the time after PCR-testing or after hospital admission between the groups. Nevertheless, it needs to be considered that the time point of infection in our cohort is, as in most similar studies, not known. The higher reactivity of T cells of critically ill patients might therefore mirror a longer course of response against the infection. Another explanation might be that the higher magnitude and functionality of the T-cell response observed in critical COVID-19 cases might simply reflect a more severe infection course with a stronger immunogenic environment, provided by a higher viral burden and inflammatory bystander activation.

Independently of the reason for the higher magnitude and functionality of CD4+ and CD8^+^ T cells in critical patients, our data demonstrate the ability of critical COVID-19 patients to mount a sufficient cellular immunity. Although the observed TH1-dominanted polyfunctional cells are commonly regarded as a parameter for protective immunity^17^,^19^,^24^, they can also provide immune damage contributing to immunopathogenesis^25^. In this context, our finding on advanced differentiation stage of SARS-CoV-2-reactive T cells found at early time points raises the question about the beneficial effect of pre-existing immunity for the course of infection. One could speculate that even though it appears to be generally protective, preexisting SARS-CoV-2 reactive T-cells with effector phenotype, which are cross-reactive with common cold corona viruses^10^, in severe infection can lead to hyperactive response and immunopathogenesis.

Although further studies are required to explore the pathogenesis of COVID-19 progression in more details, our study enhances the understanding about the complex mechanisms of the anti-SARS-COV-2 immunity that should be considered for diagnostic test and vaccine development.

## Methods

### Study population and design

This investigation is an analysis of a sub-cohort of a larger study that was published earlier ^4^. We recruited 28 patients with moderate (n=8), severe (n=10) and critical (n=10) COVID-19. The degree of COVID-19 severity was evaluated according to the guidelines of the Robert Koch Institute, Germany, as previously described^4^. The study was approved by the ethical committee of the Ruhr-University Bochum (20–6886) and University Hospital Essen (20–9214–BO), and written informed consent was obtained from all participants. The clinical and demographic patient parameters are shown in Tables S1.

Patients with moderate and severe COVID-19 were recruited after the first symptoms were reported and a positive SARS-CoV-2 PCR confirmed the disease (in median 4 days after the diagnostic test). The sampling time is presented in Table S2. For the main figures, all acquired patient samples were analyzed. For Fig.S4,only the last sample of each patient was analyzed. Samples were grouped into COVID-19 severity groups according to the symptom presentation at time of sampling

### Preparation of PBMCs and stimulation with SARS-CoV-2 overlapping peptide pools

SARS-CoV-2 PepTivator peptide pools (Miltenyi Biotec), consisting mainly of 15-mer sequences with 11 amino acids (aa) overlap containing overlapping peptide pools (OPP) spanning parts of the S-protein, or, covering the complete sequence of the N- and M-protein, were used for peripheral blood mononuclear cells (PBMC) stimulation. PBMC were prepared from EDTA collection tubes (Sarstedt) by gradient centrifugation. 2.5×10^6^ PBMC were stimulated with 1 μg/mL OPP for 16h in RPMI (Life Technologies) supplemented with 1% Penicillin-Streptomycin-Glutamin (Sigma Aldrich) and 10% FCS (PAN-Biotech). Negative controls were left untreated. Brefeldin A (1μg/ml, Sigma Aldrich) was added after 2h.

### Flow cytometry

PBMC stimulated with SARS-CoV-2 OPP were surface stained with CCR7-PerCP-Cy5.5 (clone G043H7) (BioLegend), CD4-A700 (clone OKT4) (BioLegend), Fixable Viability Dye eFluor780 (eBioscience), CD8-V500 (clone RPA-T8) (BD Biosciences) and CD45RA-BV605 (clone HI100) (BioLegend). After fixation and permeabilization (Intracellular Fixation & Permeabilization Buffer Set by Thermo Fisher Scientific) the cells were stained with Granzyme B-FITC (clone GB11) (BioLegend), IL2-PE (clone MQ1-17H12) (BioLegend), IL4-PE-Dazzle594 (clone MP4-25D2) (BioLegend), CD137 (4-1BB)-PE-Cy7 (clone 4B4-1) (BioLegend), CD154 (CD40L)-A647; (clone 24-31) (BioLegend), TNFα-eFluor450 (clone MAb11) (eBioscience), IFNg-BV650 (clone 4S.B3) (BioLegend), CD3-BV785 (clone OKT3) (BioLegend). All samples were acquired on a CytoFlex flow cytometer (Beckman Coulter).

### Data analysis and graphical representation

Flow cytometry data were analyzed using FlowJo version 10.6.2 (BD Biosciences). Gating strategies are presented in supplementary Fig. S2. Unspecific activation in unstimulated controls was subtracted from stimulated samples to account for specific activation. Antigen-specific responses above 0.001% were considered positive. Negative values were set to zero. Multifunctional T cells were analyzed using Boolean gating of IL2, IL4, IFNγ, TNFα and GrzB producing CD4^+^ and CD8^+^ T cells. Statistical analysis was performed using R, version 3.6.2^26^and GraphPad Prism v7, which was also used for graphical representation. Venn diagrams were prepared using Venny v2.1^27^.

Non-parametric statistical tests were used where applicable. Patient age and time point of sampling were compared with Kruskal-Wallis-Test. Patient gender was compared with Fisher’s exact test. Differences in T cells responses of all patient samples together were analyzed with Friedman test and Dunn’s multiple comparison test. As sphericity was not assumed, Geisser-Greenhouse correction was applied. Correlation between the T cell responses towards the different peptides was analyzed by Spearman’s rank correlation coefficient. Differences in T cell responses of patient samples grouped according to COVID-19 severity were analyzed by repeated-measurement two-way ANOVA and Tukey’s multiple comparison test. Bivariate regression analysis was performed to exclude potential bias in the analysis due to the unbalanced gender distribution of the groups. Thus, for factors significantly associated with illness severity, regression analysis was performed with gender and COVID-19 severity as independent variables, without interactions. P values below 0.05 were considered significant; only significant P values are reported.

## Data Availability

The raw data supporting the conclusions of this manuscript will be made available by the authors, without undue reservation, to any qualified researcher.

## Acknowledgments

We feel deep gratitude to the patients who donated their blood samples and clinical data for this project. We would like to acknowledge the excellent technical assistance as well as the expertise of immune diagnostic laboratory (Sarah Skrzypczyk, Eva Kohut, Julia Kurek, Jan Zapka) of Center for Translational Medicine at Marien Hospital Herne. This work was supported by grants of Mercator Foundation, the BMBF e:KID (01ZX1612A), and BMBF NoChro (FKZ 13GW0338B).

